# On the numbers of infected and deceased in the second Corona wave

**DOI:** 10.1101/2020.08.10.20171553

**Authors:** Rainer Janssen, Juergen Mimkes

## Abstract

**Summary:** In Germany and other countries, a second wave of corona infections has been observed since July 2020, after the first wave has subsided. We have investigated both waves by a modified SIR-SI infection model, adapted to the data to the Robert-Koch-Institute (RKI) or the Johns-Hopkins-University (JHU).

The first wave is characterized by the SIR model: in a perfect lockdown only a small part of the society is infected and the infections end after a certain time. The SI part considers the incompleteness of any lockdown: at the end of the first wave infections do not completely go down to zero, but continue to rise again, but only slowly due to mouth protection, hygiene and distance keeping. During this first wave the number of deceased people follows the number of infected persons with a fixed time interval and percentage: mostly symptomatic ill people have been tested. This applied to nearly all countries observed, with different intervals and percentages.

In the present second wave, the number of daily infections has risen again significantly in some countries, and it may be questioned whether this is due to the increased number of tests. The answer may be given by looking at the daily number of deaths. In Germany, Austria, Italy, Great Britain and others this number has still remained at a constant level for six weeks. In these countries a second wave of died people has not yet arrived. The increased number of tests include obviously mostly asymptomatically infected persons, who do not fall ill or die from coronavirus. However, in some countries, like USA or Israel, the second wave did arrive. The numbers of infected and deceased people both have grown. A real second wave is a permanent threat to all countries.

## Introduction

Since February 2020, Germany and other countries have been affected by the coronavirus pandemic. A pandemic normally affects the entire population, but the population is often able to protect itself to a large extent through immunity or vaccination. Since this protection does not yet exist in the coronavirus pandemic, almost all affected countries have ordered a temporary lockdown and restriction of contacts in order to keep the number of seriously ill and dead people as low as possible.

The SIR model of Kermack and McKendrick [1] is used to describe the course of the pandemic with time. Many authors apply this model to analyze and predict the course of the coronavirus pandemic in Germany and other countries [2–6] using data from the Robert-Koch-Institute (RKI) and the Johns-Hopkins-University (JHU). The SIR model represents the mathematical relationship between the three groups of an infection: the contagious (S, susceptible), the infectious (I) and the convalescent (R). The last group includes the deceased. The sum of the three groups, S + I + R = N, gives the fixed number N of the infectable people. Sometimes the simpler analytical solution of the SI model is used, neglecting the group of convalescents (R). This SI-model generally describes the course of infection with sufficient accuracy [7].

In this paper, the infection rates of the first and second wave in Germany will be examined in detail and compared to other countries.

## Data of the corona pandemic in Germany

Figure 1a shows the course of corona infections and deaths in Germany from February to August 2020 according to data from the RKI. The course of the infections in logarithmic representation in Figure 1a shows that between February and April the number of infections increases exponentially. However, not all 83 million inhabitants in Germany are infected, only about 200,000 people. This limitation is due to contact restrictions. The slight increase in infections from April onwards shows that the first wave is not finished and some people continue to become infected through gaps in the contact ban. But the increase in infections between April and July is much lower than between February and April. Until July the number of persons deceased with the coronavirus follow the number of infected persons by 13 days with a percentage of 4.8 %. This enables us to calculate an expected number of deceased persons about 13 days ahead. We observe similar trends in Italy (5 days and 14.6%), Austria (13 days and 4.1%), the Great Britain (4 days and 15.5%), Israel (11 days and 1.8%), Japan (11 days and 5.5%) or the USA (6 days and 7.2%).

**Figure 1a.**
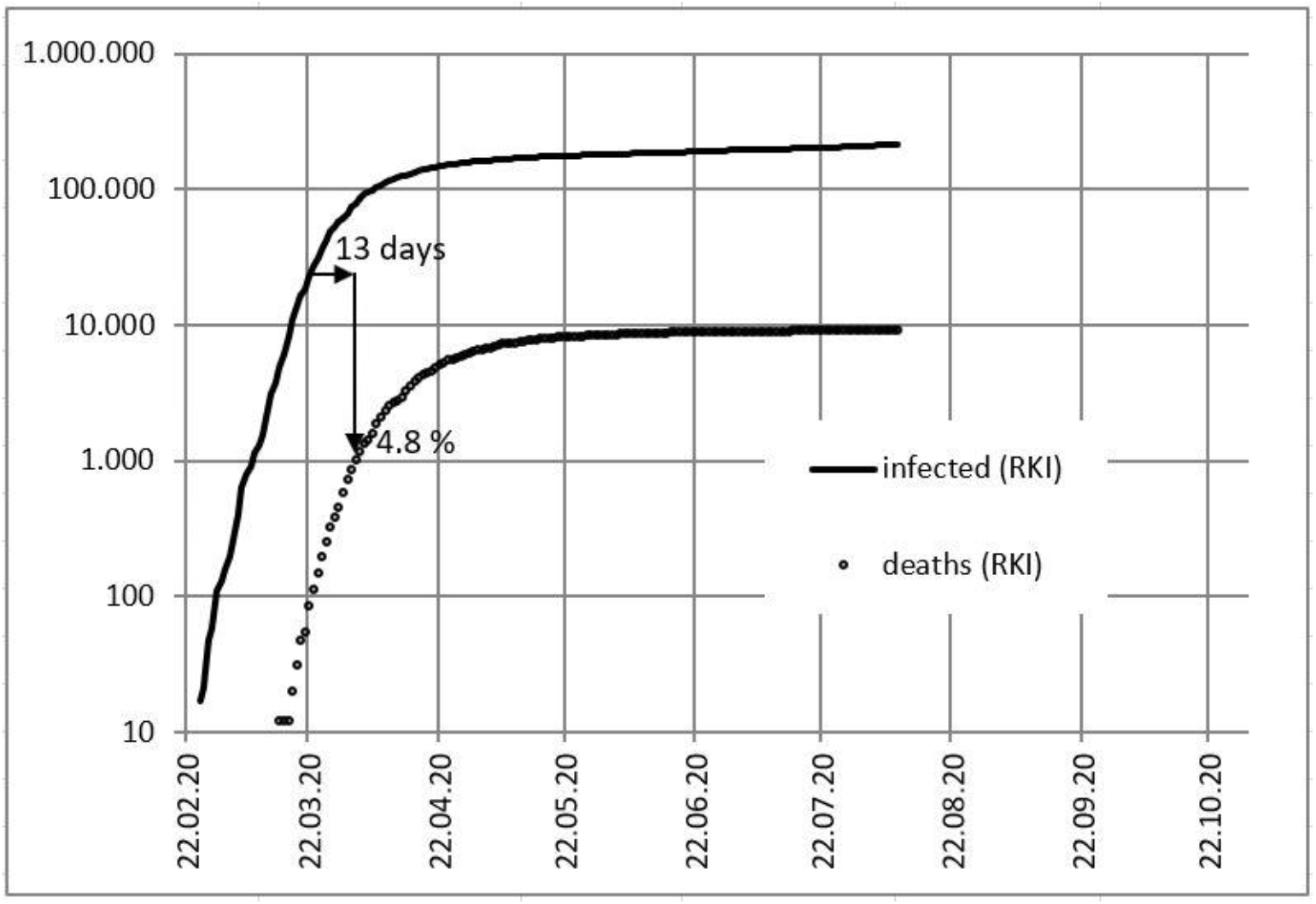
Logarithmic representation of confirmed infections in Germany and of those who died with the coronavirus from February to August 2020 according to data from the RKI.

## A modified SIR-SI model of infections in the first wave in Germany

The more precise course of the infections can be seen in the linear representation of the infections. In Figure 1b we have adjusted the data of the infections up to April to the SIR model. According to this model, about 160,000 people will be infected with the coronavirus during the contact ban and lockdown until the end of May. Later on, there should be no new infections. But already from April onwards it became apparent that the contact restrictions were incomplete, 200,000 people were infected by mid-July.

**Figure 1b.**
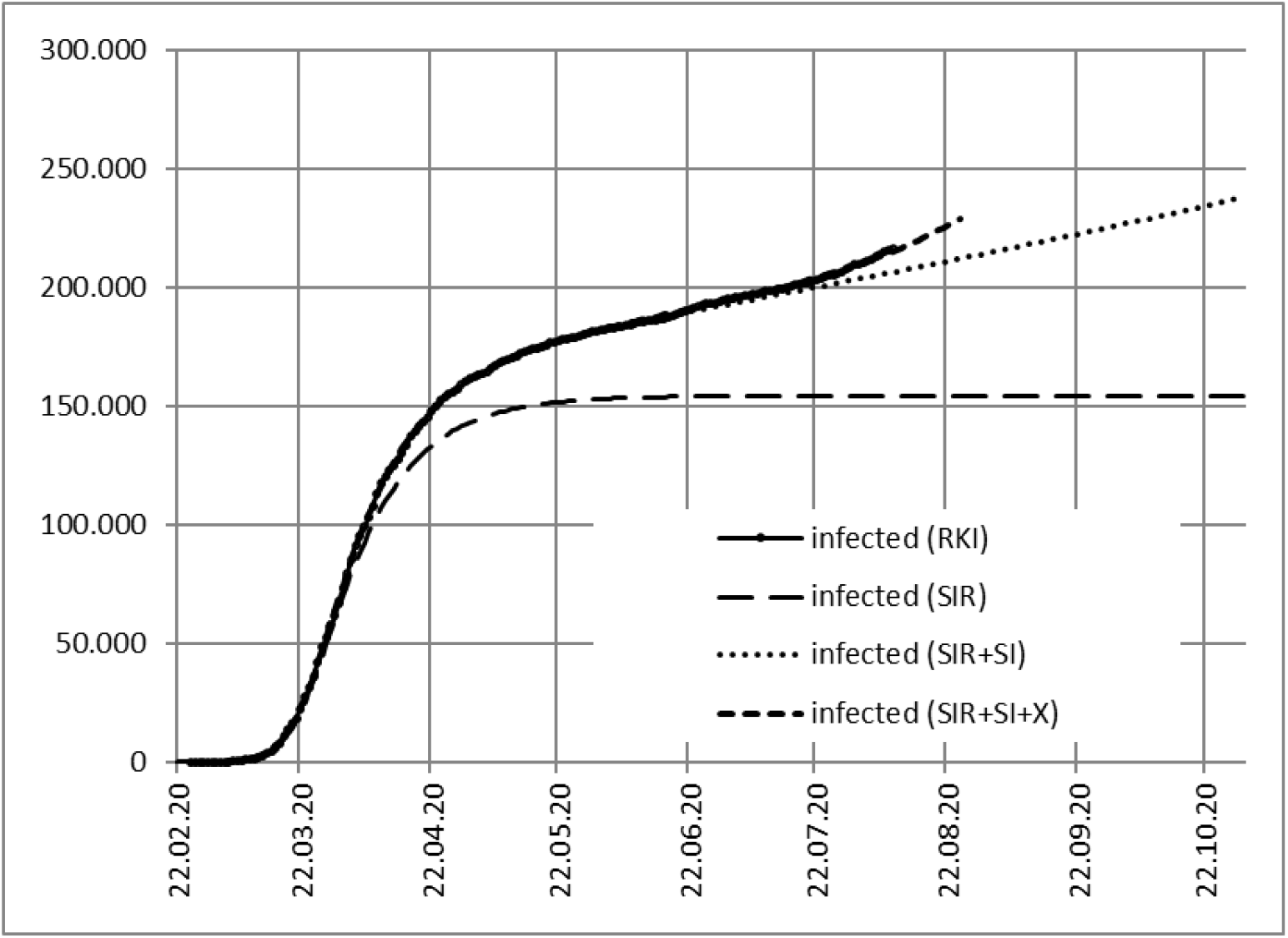
Number of confirmed infections in Germany from February to August 2020 and the adaptation of the SIR-SI-model to the data of the first wave according to RKI.

As the SIR model does not reflect the full course of the first wave, we have coupled the SIR-model with an SI-model that simulates the low probability of infections due to the incompleteness of the lockdown from the beginning of the pandemic. The probability of infection in the SI-model at the end of the first wave is only about 0.2 %, due to mouth protection, hygiene and distance keeping. Accordingly, centers of infection have not spread to the population. Only from July 2020 on we find a deviation (SIR-SI-X) from our SIR-SI-model of the first wave, indicating a second wave.

Figure 1c shows the first wave of the daily new infected persons in Germany. The combined SIR-SI-model has been adjusted to the data according to RKI. Only from mid-July on a second wave seems to be growing (SIR-SI-X).

**Figure 1c.**
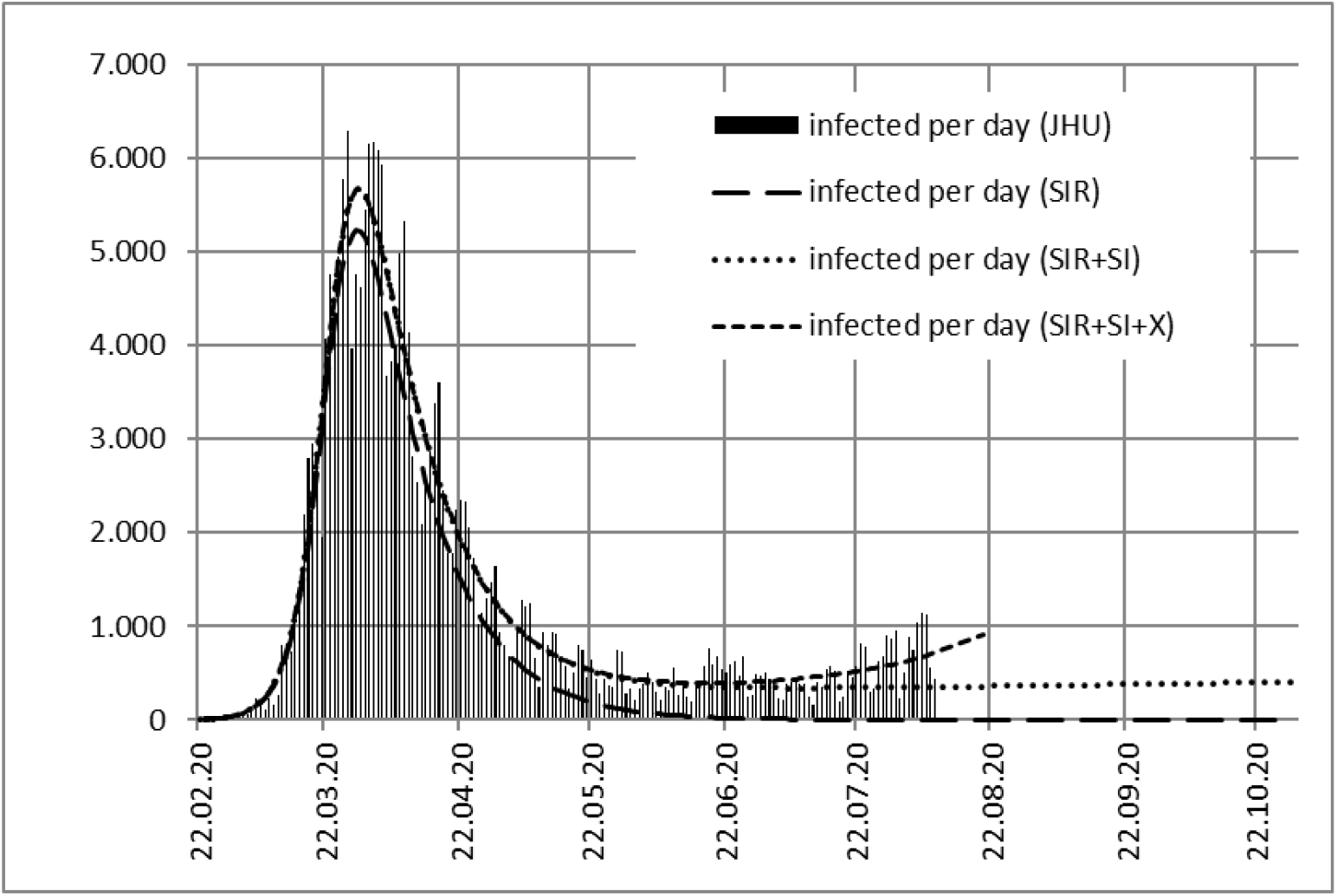
Number of confirmed daily new infections in Germany from February until August 2020 and the adaptation of the SIR-SI-model to the data of the first wave according to RKI.

## Number of deceased persons in the pseudo second wave in Germany

From July onwards, Figure 1b and Figure 1c indicate a rise of infection numbers (SIR-SI-X) that deviates from the new SIR-SI-model of the first wave. This is discussed in Figures 1d and 1e.

**Figure 1d.**
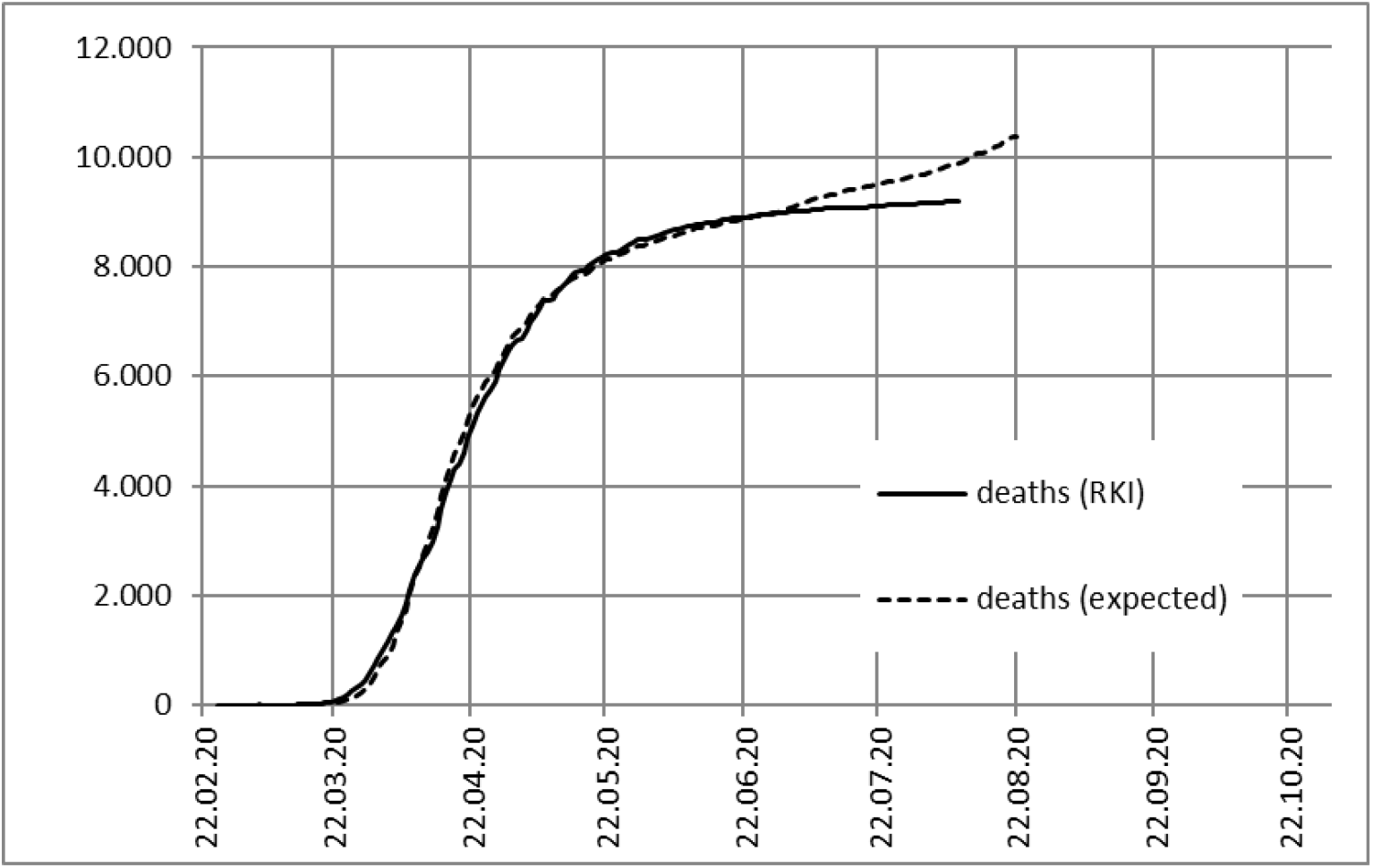
Number of reported deaths in Germany compared to the expected number, not indicating a real second wave of deaths.

**Figure 1e.**
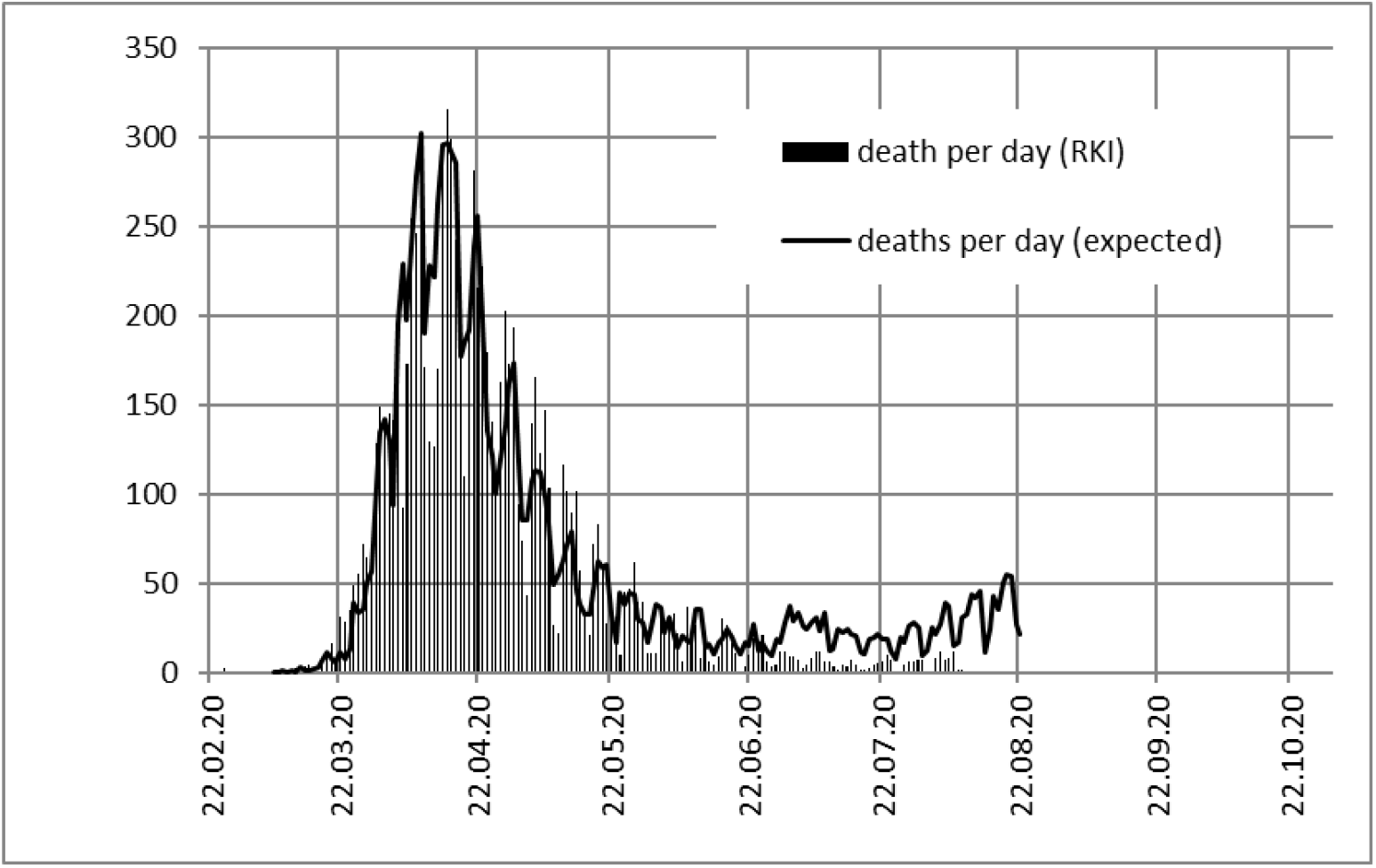
Number of deaths reported daily in Germany compared to the expected number, not indicating a real second wave of deaths.

Figure 1d shows the course of accumulated deceased persons (solid line) compared to the expected number of deceased (dashed line): until July 2020 both lines agree in Germany with a time lag of 13 days and a percentage of about 4.8% (see Figure 1a). But from July onwards, Figure 1d indicates constant numbers of deceased that deviate from the expected numbers.

Figure 1e shows the course of daily deceased persons (bars) compared to the expected number of deceased (solid line): until July 2020 both lines agree in Germany, but since then less people are dying than expected from the number of infections. Figure 1e shows that although a “second wave” of infected persons is indicated, the number of daily reported deaths remains at a low level, so that a second wave of the corona pandemic is currently not apparent in Germany. A similar phenomenon is also observed in Austria, Italy, Great Britain or Japan.

## Number of deceased persons in real second waves in USA and Israel

Figure 2a shows the number of daily new infections in USA, which has risen sharply in a second wave since June. The wave seems to follow an SI model but has been called X here unless it is verified.

**Figure 2a.**
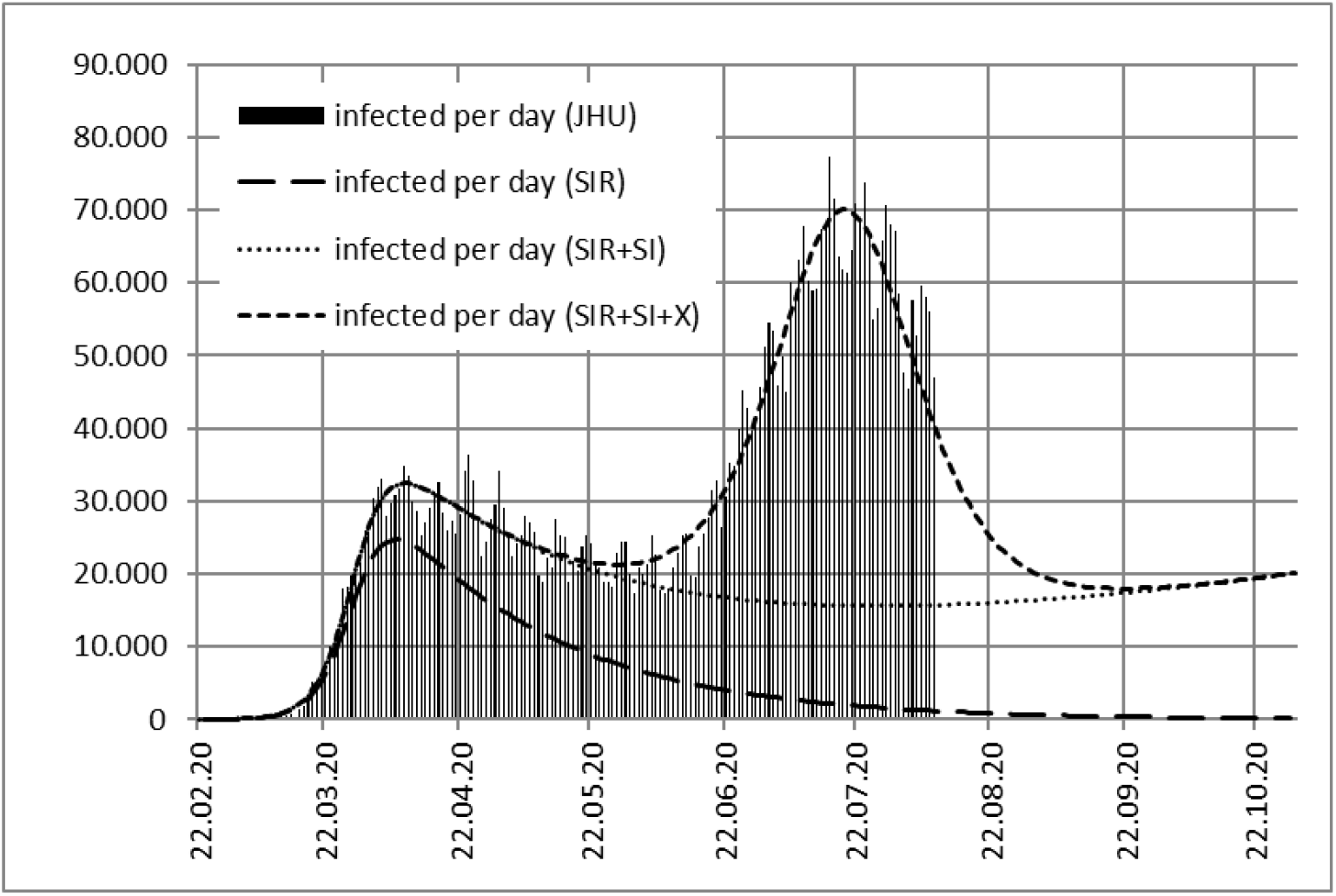
Number of new infections in the USA and their modelling.

Figure 2b presents the second wave of daily expected deceased in USA compared to the reported number of daily deceased persons. The growing number of tests have raised the number of infections. The number of daily reported deceased has not grown as much as would be expected from infections. But the course of daily reported deceased clearly indicates a second wave in USA since June 2020.

**Figure 2b.**
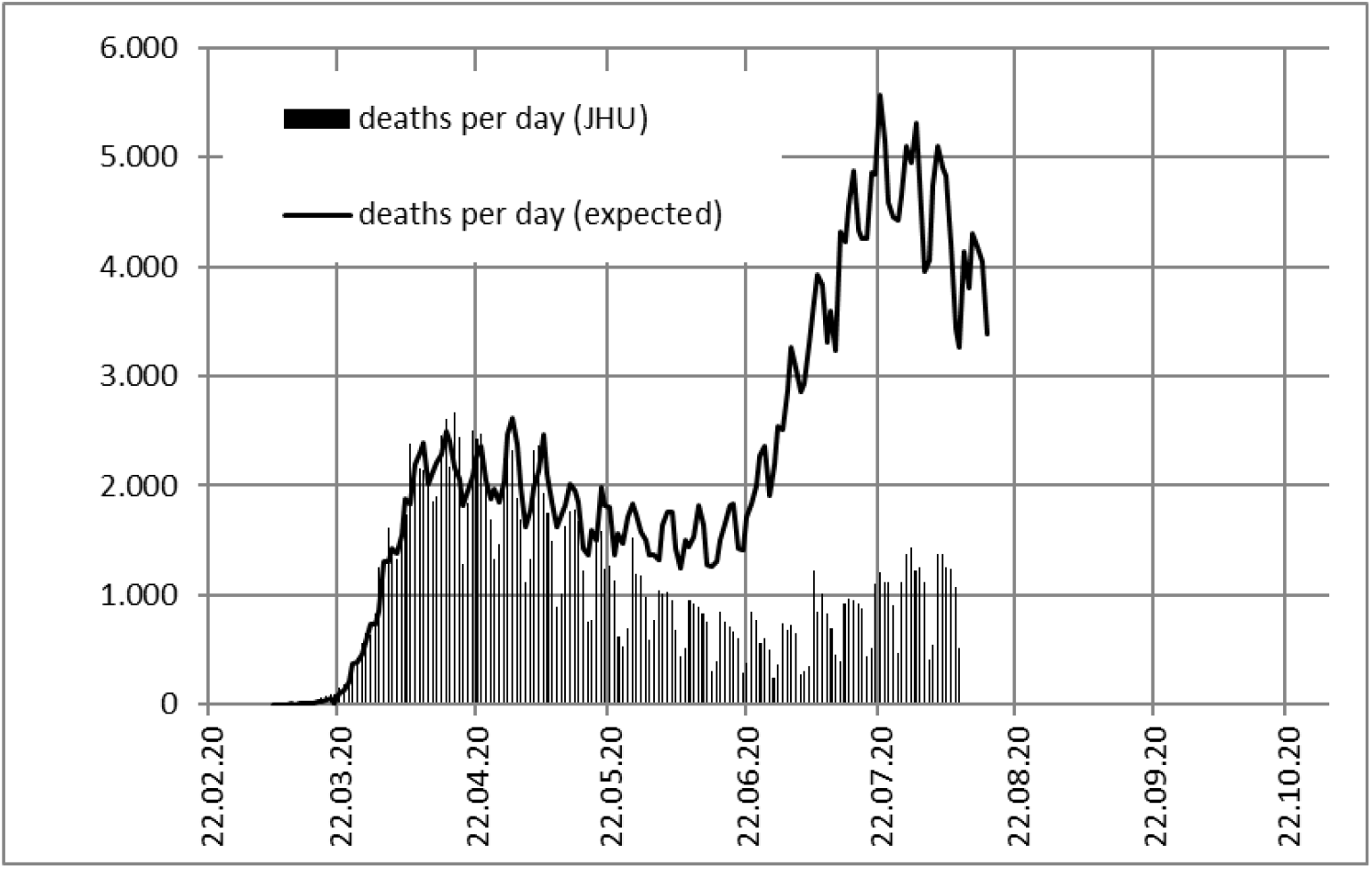
Number of deaths reported daily in the USA compared to the expected number. The growing number of daily reported deceased clearly indicates a real second wave.

Figure 2c shows another real second wave in Israel. Obviously, the second wave in Israel is still of the same size as the first wave. Due to more testing considerably more asymptomatic cases were recorded in the second wave.

**Figure 2c.**
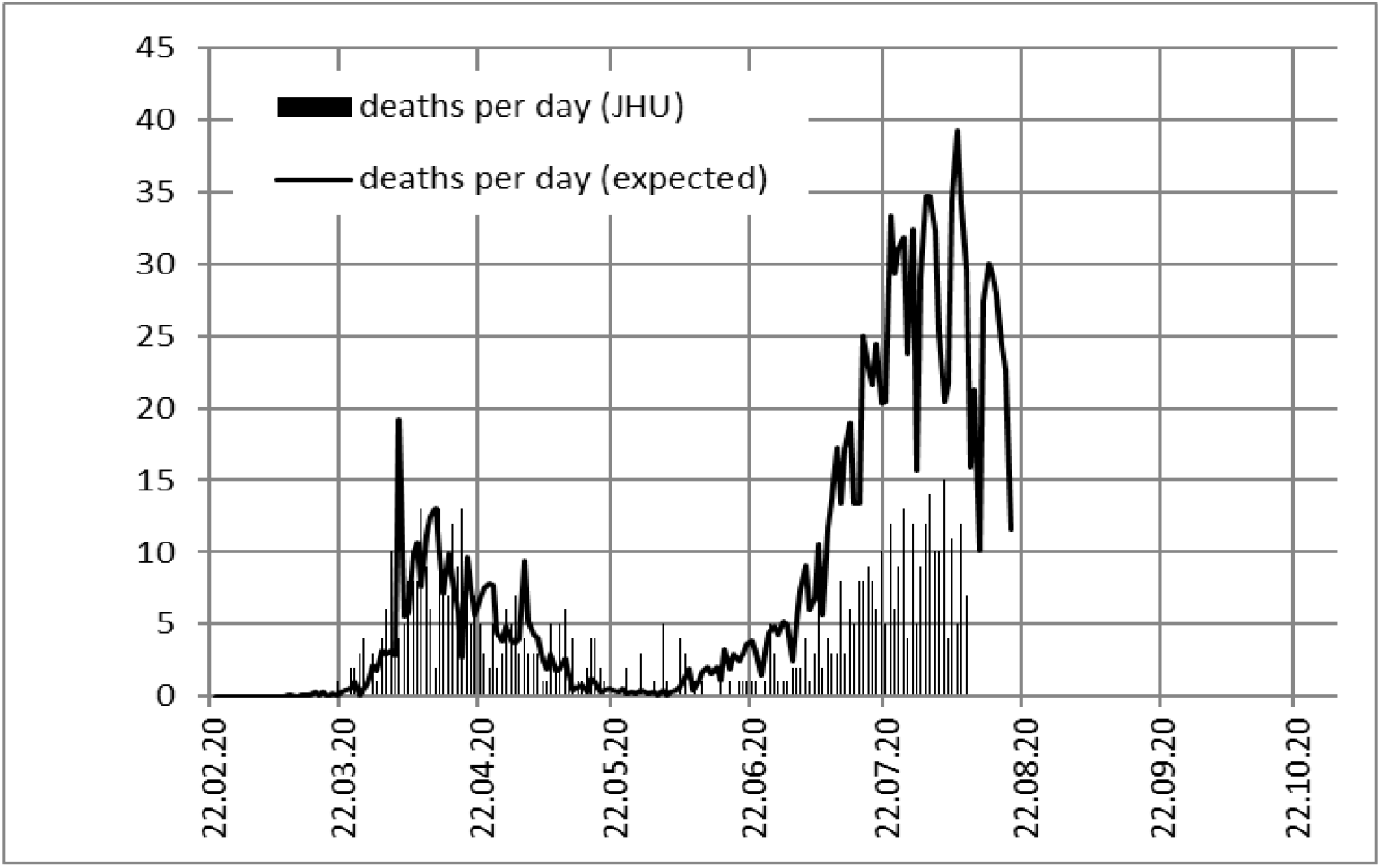
Number of deaths reported daily in Israel compared to the expected number. The growing number of daily reported deceased clearly indicates a real second wave.

## Conclusion

In all the countries the numbers of daily reported infections were most important to understand the first wave. Presently, the numbers of daily reported deaths are most important to understand the second wave. Nevertheless, it remains necessary to carry on many meaningful tests to prevent potentially infectious persons from passing on infections. A second wave still may come to all countries.

In the future the authors plan to make publicly available the used Excel files with the data of all countries in an updated form: https://physik.uni-paderborn.de/alumni/mimkes/

## Data Availability

JHU and RKI corona data files

